# Deep Learning Image Analysis of Benign Breast Disease to Identify Subsequent Risk of Breast Cancer

**DOI:** 10.1101/2020.09.02.20186643

**Authors:** Adithya D Vellal, Korsuk Sirinukunwattana, Kevin H Kensler, Gabrielle M Baker, Andreea L Stancu, Michael E Pyle, Laura C Collins, Stuart J Schnitt, James L Connolly, Mitko Veta, A Heather Eliassen, Rulla M Tamimi, Yujing J Heng

## Abstract

**Background:** New biomarkers of risk may improve breast cancer risk prediction. We developed a computational pathology method to segment benign breast disease (BBD) whole slide images (WSIs) into epithelium, fibrous stroma, and fat. We applied our method to the BBD breast cancer nested case-control study within the Nurses’ Health Studies to assess whether computer-derived tissue composition or a morphometric signature was associated with subsequent risk of breast cancer.

**Methods:** Tissue segmentation and nuclei detection deep-learning networks were established and applied to 3795 WSIs from 293 cases who developed breast cancer and 1132 controls who did not. Percentages of each tissue region were calculated and 615 morphometric features were extracted. Elastic net regression was used to create a breast cancer morphometric signature. Associations between breast cancer risk factors and age-adjusted tissue composition among controls were assessed using analysis of covariance. Unconditional logistic regression, adjusting for the matching factors, BBD histological subtypes, parity, menopausal status, and BMI evaluated the relationship between tissue composition and breast cancer risk.

**Results:** Among controls, BBD subtypes, parity, and number of births were differentially associated with all three tissue regions (*p*< 0.05); select regions were associated with childhood body size, BMI, age of menarche, and menopausal status (*p*< 0.05). Higher proportion of epithelial tissue was associated with increased breast cancer risk (OR = 1.39, 95% CI 0.91–2.14 comparing highest and lowest quartiles; *p-trend< 0.05)*. No morphometric signature was associated with breast cancer.

**Conclusion:** The amount of epithelial tissue may be incorporated into risk assessment models to improve breast cancer risk prediction.

## Introduction

Breast cancer is the most common cancer in women. One in eight women in the United States will develop breast cancer in her lifetime^1^ Whilst early detection is imperative, identifying and lowering breast cancer risk may help reduce breast cancer morbidity and mortality. Breast cancer risk factors may be non-modifiable (e.g., genetics, dense breast tissue, and benign breast disease (BBD)) or modifiable (e.g., adiposity and alcohol consumption). Researchers continue to identify new biomarkers of risk^2–7^ as well as update risk assessment models^8–12^ to improve breast cancer risk prediction.

Technological advances have enabled the engineering of deep-learning algorithms to analyze whole slide images (WSIs) for disease detection and diagnosis^13–18^, including discriminating between breast cancer and benign breast tissue^19–21^. Our group recently developed a novel deep-learning based computational pathology method to capture quantitative measures of breast terminal duct lobular unit (TDLU) involution^22,23^, a purported biomarker of breast cancer. We reported that neither quantitative measures of TDLU involution derived by our technology nor traditional manual qualitative assessment by pathologists was associated with breast cancer among women diagnosed with BBD^24^.

In this manuscript, we engineered another deep-learning based computational pathology method to 1) segment BBD histopathological images into epithelial, fibrous stroma, and fat regions; 2) calculate the amount of each tissue region expressed as a percentage of total tissue; and 3) extract morphometric features from each tissue region. We applied our method to the BBD breast cancer nested case-control study within the Nurses’ Health Study (NHS) and NHSII to evaluate whether computer-derived tissue composition or a morphometric signature in women diagnosed with BBD was associated with subsequent risk of breast cancer.

## Materials and Methods

### Study population

The Nurses’ Health Study (NHS) and NHSII were established in 1976 and 1989, with 121,700 US female registered nurses aged 30–55 years and 116,429 nurses aged 25–42 years, respectively. NHS/NHSII participants first completed baseline questionnaires that provided a medical history as well as extensive information about demographic, lifestyle, reproductive, and dietary risk factors for breast cancer^25^. In biennial follow-up questionnaires, participants provide updated information and report new diagnoses of BBD or breast cancer. Participants who reported a diagnosis of BBD were contacted for consent to retrieve pathology records and BBD lesion specimens for centralized pathology review^4,26,27^. Participants who reported breast cancer were confirmed verbally by the participant, via medical record review, or via the cancer registry. Eligible women with biopsy-confirmed BBD were placed into two sub-studies within the NHS/NHSII—the BBD Incidence study^28–31^ and/or the BBD breast cancer nested case-control (NCC) study^2–4,24,26,27,32–35^. whole slide images (WSIs) from women in the BBD Incidence study were used in the development phase to engineer the deep-learning networks. The BBD NCC study was used in the application phase to determine if computer-derived tissue composition or morphometric features were associated with subsequent breast cancer.

### Development Phase: Engineering the tissue segmentation network

Forty-eight hematoxylin and eosin (H&E) histopathological slides from the BBD Incidence study were selected and digitized at 40x magnification (Panoramic SCAN 150, 3DHISTECH Ltd, Budapest, Hungary). These 48 WSIs encompassed a wide variety of epithelial morphology from normal epithelium (i.e., TDLUs; n = 6)) to non-malignant BBD lesions classified into three histopathological subtypes. The non-proliferative subtype was represented by cysts (n = 5) and fibroadenoma (n = 5). Proliferative without atypia subtype included intraductal papilloma (n = 5), radial scars (n = 5), adenosis (n = 5), columnar cell change (n = 4), and usual ductal hyperplasia (n = 5). Proliferative with atypia lesions consisted of atypical ductal hyperplasia (n = 2), atypical lobular hyperplasia (n = 4), and flat epithelial atypia (n = 2). For each WSI, four patches of 2048 × 2048 pixels were selected with at least one patch containing the lesion. Each patch was first manually annotated by a researcher for epithelium, fibrous stroma, fat, and background, and later manually verified by a board-certified pathologist (Supplementary Figure 1). The 48 WSIs were split into training and testing sets in a 3:1 ratio. Supplementary Figure 2A displays examples of patches used to train the network.

To segment BBD images into background, epithelial (normal TDLUs, TDLUs exhibiting proliferative or metaplastic changes, and various BBD lesions), fibrous stroma (inter- and intralobular), and fat regions, we engineered a custom 21-layer fully convolutional network inspired by VGG-Net^36^ and U-Net^37^ (Supplementary Table 1). This network operated at a magnification level of 2.5x, using the sliding window approach^38^ with an input size of 128 × 128 pixels to perform segmentation. To ensure that the network focused on differentiating tissue types by texture and was indifferent to variations in staining, data augmentation in the form of color normalization, random color and intensity shifts and geometrical image transformations were performed during training. Patches were flipped vertically and horizontally, transposed, and altered in hue, brightness, contrast, and saturation prior to every iteration.

Training was completed in two steps, using an active learning process^39^. The network was first trained using a complete training set consisting of over 35,000 patches. Uncertainty measure was computed for each training patch through pixel-wise margin sampling^40^. The 7,500 patches with the highest certainty and the 7,500 patches with the lowest certainty were then used to construct a smaller training set. This new 15,000 patch training set was used to retrain the network. This training methodology was utilized in order to prevent the inclusion of ambiguously annotated patches in the training set. The log-likelihood function was used as a loss measure. In order to mitigate class imbalance, each term of the loss function was normalized based on the distribution of the corresponding tissue type at a patch-by-patch level. Additionally, each class was weighted with empirically determined coefficients to achieve the best overall model performance based on the test set. The background weight was 0.7, epithelium weight was 0.9, fibrous stroma weight was 1.4, and fat weight was 1.1. Adaptive moment estimation optimization^41^ with an initial learning rate of 10^−4^ and batch size of eight was used throughout training.

### Development Phase: Engineering the cell nuclei detection network

One of the goals of this computational method was to extract morphometric features from tissue regions. Some of these morphometric features require additional information from cell nuclei. Thus, a nuclei detection network was also created using a set of 30 H&E breast cancer WSIs from The Cancer Genome Atlas where cell nuclei had been previously annotated^42^. Nuclei detection was performed using a fully convolutional U-Net architecture^37^. Images were segmented into nucleus, nuclei membrane, and background, with the nuclei membrane label being established to ensure proper separation of overlapping nuclei. Any pixel labeled as nuclei membrane by the network was re-classified as part of the background before generating the final binary nuclei mask. This network segmented nuclei at 10x magnification and used the sliding window approach^38^, with an input size of 144 × 144 pixels for segmentation. While no two-stage training scheme was employed with this model, the details of the color normalization, data augmentation, training, and loss function steps were similar to the tissue segmentation network. The two notable differences were that the log-likelihood loss function was not additionally weighted by class, and each training batch consisted of 32 patches rather than eight. See Supplementary Figure 2B for example patches used for training this network. All networks were developed using the Tensorflow 0.12.0 API for Python 2.7. The source code for our networks is available at https://github.com/avellal14/BBD_Pipeline.

### Development Phase: Statistical Analysis

To evaluate the tissue segmentation network, precision, recall, and Dice similarity coefficient were calculated using the held-out test set (n = 48). Dice similarity coefficient is the harmonic mean of precision (i.e., sensitivity) and recall (i.e., positive predictive value) and assesses how accurate the automated segmentation compares with ground truth on a pixel-wise basis. The range for Dice similarity coefficient is from 0 to 1, with 1 indicating perfect overlap. For nuclei detection, three-fold cross validation was used to obtain the precision, recall, and Dice similarity coefficient of the network.

### Application Phase: BBD NCC study participants

The study protocol was approved by the institutional review boards of the Brigham and Women’s Hospital and Harvard T.H. Chan School of Public Health, and those of participating registries as required. The BBD NCC study consisted of 293 cases and 1132 controls (total n = 1425). Cases reported a diagnosis of invasive breast cancer after the cohort baseline (through 1998 for NHS, through 1999 for NHSII) and had previously reported a BBD diagnosis (either prior to study entry or after study baseline); were diagnosed with breast cancer a median of 7.67 years after BBD diagnoses (interquartile range 4.33 to 11.75 years); and were excluded if the time between BBD and breast cancer diagnoses was less than six months or if there was evidence of invasive or *in situ* carcinoma during centralized histopathological review of the BBD lesion. Tumor estrogen receptor (ER) status was obtained from centralized review of breast tissue microarrays^43^. If centralized review data was unavailable then ER status was retrieved from pathology reports. Controls were women diagnosed with BBD who did not develop breast cancer. Cases and controls were matched 1:4 on year of BBD diagnosis, age at breast cancer diagnosis (index date for controls), and years between BBD and breast cancer diagnosis (or index date). A total of 3795 slides were digitized at 20x (n = 213) or 40x magnification (n = 3582). Each woman contributed between one to four WSIs (median WSIs n = 3).

The histological type of the BBD lesion was determined by central pathology review. Participant body mass index (BMI), age at menarche, parity, age at first birth, breastfeeding history, and menopausal status were obtained from questionnaires of the participants closest to but prior to BBD biopsy. The average body sizes at ages 5 and 10 reported by using a nine-level pictogram (Level 1 as leanest)^34^. Birth index, a surrogate metric that reflects the timing and spacing of births, was calculated as previously described^44^. A higher birth index indicates a higher number of births occurring at earlier ages.

### Application Phase: Applying our networks to WSIs from the BBD NCC study participants

Figure 1 shows an overview of our BBD image analysis pipeline. For each WSI, binary tissue masks were generated using simple color thresholding at a magnification level of 0.25x (Figure 1B). After tissue-containing areas were located, each WSI was split into patches of size 2048 × 2048 pixels. Each patch was extracted at a 10x magnification level, and tissue segmentation and nuclei detection were performed using the networks established in the development phase (Figure 1C). Each patch resulted as a segmentation map with each pixel classified as epithelium, fibrous stroma, fat, or background.

**Figure 1.**
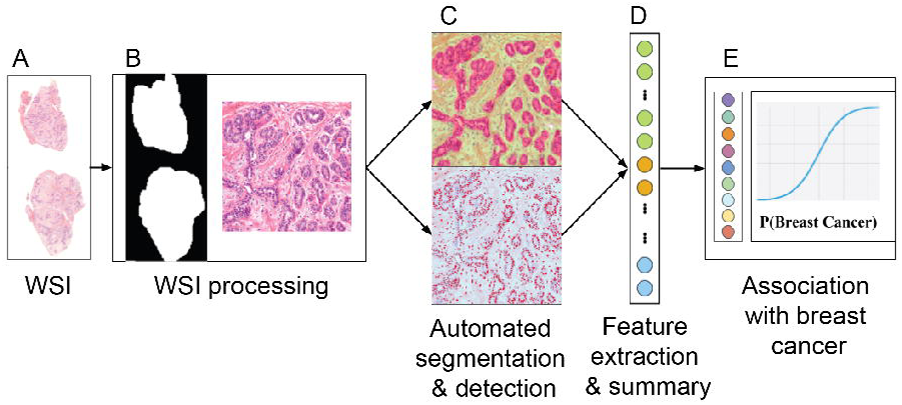
Overview of our BBD image analysis pipeline. **(A)** A whole slide image (WSI). **(B)** Image processing to extract tissue-containing areas of the WSI. **(C)** Applying our tissue segmentation and nuclei detection networks created in the development phase to a WSI to obtain a segmentation map. **(D)** From the segmentation map, computer-derived morphometric features were extracted. Percentages of tissue regions were also computed from the map. Morphometric data were summarized from all WSIs belonging to the same woman. **(E)** Identifying if morphometric features are associated with breast cancer.

### Application Phase: Extracting percentages of each tissue region

Each tissue region was expressed as a percentage of the total amount of tissue analyzed for each woman. Pixels classified as epithelium, fibrous stroma, or fat were individually summed across patches from a single WSI, combined across WSIs pertaining to each woman, and divided by the total number of pixels detected across all tissue regions.

### Application Phase: Extracting morphometric features

Morphology, texture, and graph-based spatial features (i.e., computer-derived morphometric features; n = 619) were extracted using the WSIs in conjunction with the automated tissue segmentation and nuclei detection results (Figure 1D). Since fat regions were mostly empty white spaces, fat and fibrous stroma regions were combined as stroma for feature extraction. For each tissue region, 25 morphological features summarized the areas, perimeters, spatial densities, and red green blue color distributions of detected nuclei; 161 features summarized various geometric properties (e.g. eccentricity, convex area) and image moments^45^; 113 Haralick texture^46^ and local binary pattern^47^ features were calculated using the grayscale co-occurrence matrices. The 598 features detailed above were computed at the patch level and then aggregated across all patches belonging to a single WSI. At the WSI level, an additional 21 graph-based spatial features (i.e., Voronoi diagrams^48^ and Delaunay triangulations^49^) were computed directly to characterize the holistic spatial distribution of the epithelial regions.

For women with more than one WSI, the value for each feature was further summarized using the median calculated across all her WSIs. Four features were subsequently excluded due to missing values—percentage area and the standard deviation of major axis length for both epithelial and stroma areas. Thus, a total of 615 features were analyzed in this study.

### Application Phase: Building a morphometric signature

The 1425 study participants were split into a training:test ratio of 60:40. The *sampling* package in R ensured the test set of 570 women (40%) was representative of the training set by selecting women based on outcome (case/control) and BBD histopathological subtypes. A morphometric signature associated with breast cancer was constructed using a training set of 855 women (60%), 615 z-scored morphometric features, and elastic net regularized regression model with 1000 iterations and 10-fold cross validation *(glmnet* package in R^50^). The morphometric signature was subjected to bootstrapping (1000 iterations) to obtain 95% confidence intervals for feature coefficient estimates. Features whose coefficients with 95% confidence intervals crossing zero were dropped from the signature. A signature score for each woman in the test set was computed by summing the products of the final selected features’ coefficients and z-scored values.

### Application Phase: Statistical Analysis

Preliminary assessments using Wilcoxon rank sum test evaluated if there was any difference in amount of epithelial, fibrous stroma, and fat tissues between cases and controls. Kruskal-Wallis test was used to determine if each tissue region was different between case/control statuses, stratified by BBD histological subtypes. Significant Kruskal-Wallis tests were further evaluated using Dunn’s post hoc tests with Benjamini-Hochberg multiple testing method to obtain adjusted p-values (*FSA* R package 0.8.30^51^). Only meaningful significant (adjusted *p*< 0.05) comparisons within cases, controls, and between case and controls were reported.

The associations between breast cancer risk factors and tissue composition (natural log-transformed) among controls were assessed using analysis of covariance (ANCOVA) adjusting for age at BBD biopsy *(emmeans* R package version 1.4.4^52^). Each tissue region was categorized into quartiles as defined by the distribution among controls. Unconditional logistic regression models accounting for the matching factors to estimate odd ratios (ORs) and 95% confidence intervals (CI) was used to determine the relationship between each tissue region (in quartiles) and breast cancer risk (Figure 1E). Unconditional logistic regression models were used because of incomplete matched case-control sets—lack of pathology records and/or slides for all selected cases and controls. Model 1 adjusted for matching factors (year of BBD biopsy, age at index date, time between BBD biopsy and index date); model 2 adjusted for matching factors and BBD histological subtypes; and model 3 adjusted for matching factors, BBD histological subtypes, parity, menopausal status, and BMI. Analyses were also conducted by stratifying the women according to BBD histological subtype, parity, menopausal status, or BMI. Polytomous logistic regression models assessed the association between each tissue region and risk of breast cancer defined by tumor ER expression.

The morphometric signature scores were winsorized to 96% where scores below the second percentile were set to the score at the second percentile and scores above the 98th percentile were set to the score at the 98th percentile. The association of this score and breast cancer outcome in the held-out test set was similarly evaluated using unconditional logistic regression models. The level of significance used for all statistical tests was *p*< 0.05. All statistical analyses were performed using R.

## Results

### Tissue segmentation and nuclei detection networks performance (development phase)

The precision, recall, and Dice similarity coefficient of the tissue segmentation network were 0.79, 0.71, and 0.75 for epithelium; 0.83, 0.88, and 0.85 for fibrous stroma; 0.85, 0.67, and 0.75 for fat; and 0.84, 0.92, and 0.88 for background. The cell nuclei detection network detected nuclei with a precision of 0.93, recall of 0.71, and Dice similarity coefficient of 0.81. An example of an original image, ground truth annotation, and automated segmentation or detection for each network is presented in Figure 2.

**Figure 2.**
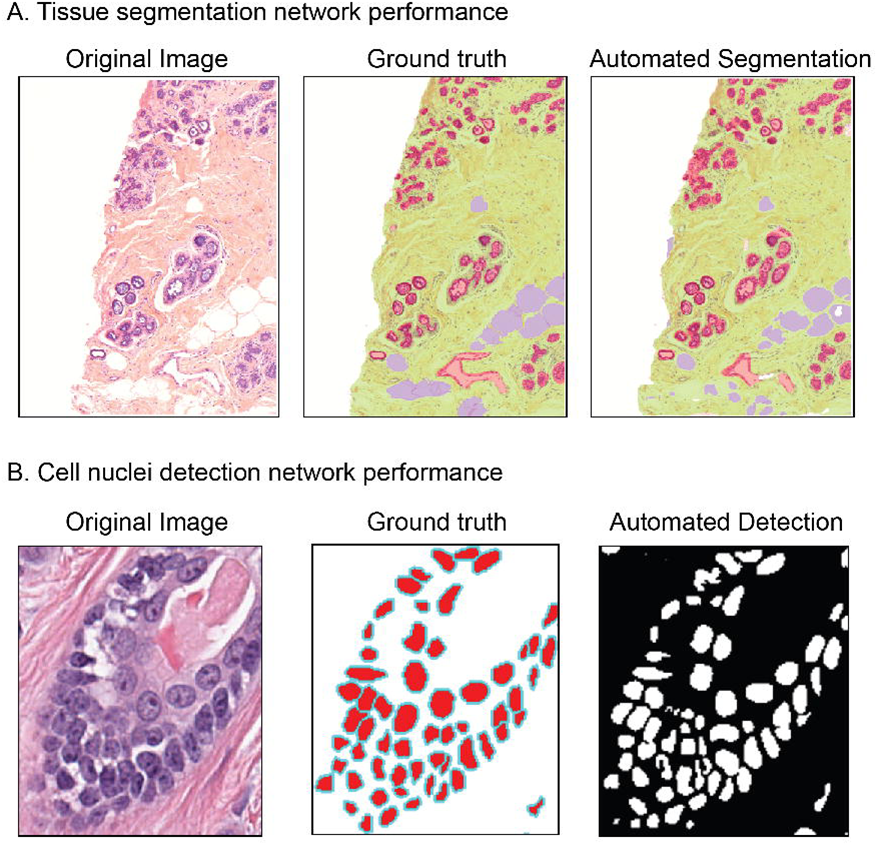
An example of an original image, ground truth, and automated segmentation or detection for each deep-learning network. **(A)** For tissue segmentation, white represents background, green represents fibrous stroma, red is epithelium, and purple is fat. **(B)** For cell nuclei detection, white represents background, red is nucleus, and cyan is nuclei membrane border. The final output produces a binary mask which considers nucleus membrane pixels to be part of the background.

### BBD NCC study participants

Table 1 displays the matching factors and BBD histopathological subtypes of the 293 breast cancer cases and 1132 controls. The majority of the women were diagnosed with proliferative breast disease without atypia. The average age at breast cancer diagnosis among cases was 54.1±8.7 standard deviation (SD). Among the 293 cases, 185 tumors were ER-positive, 51 were ER-negative, and 57 were unknown.

**Table 1.**
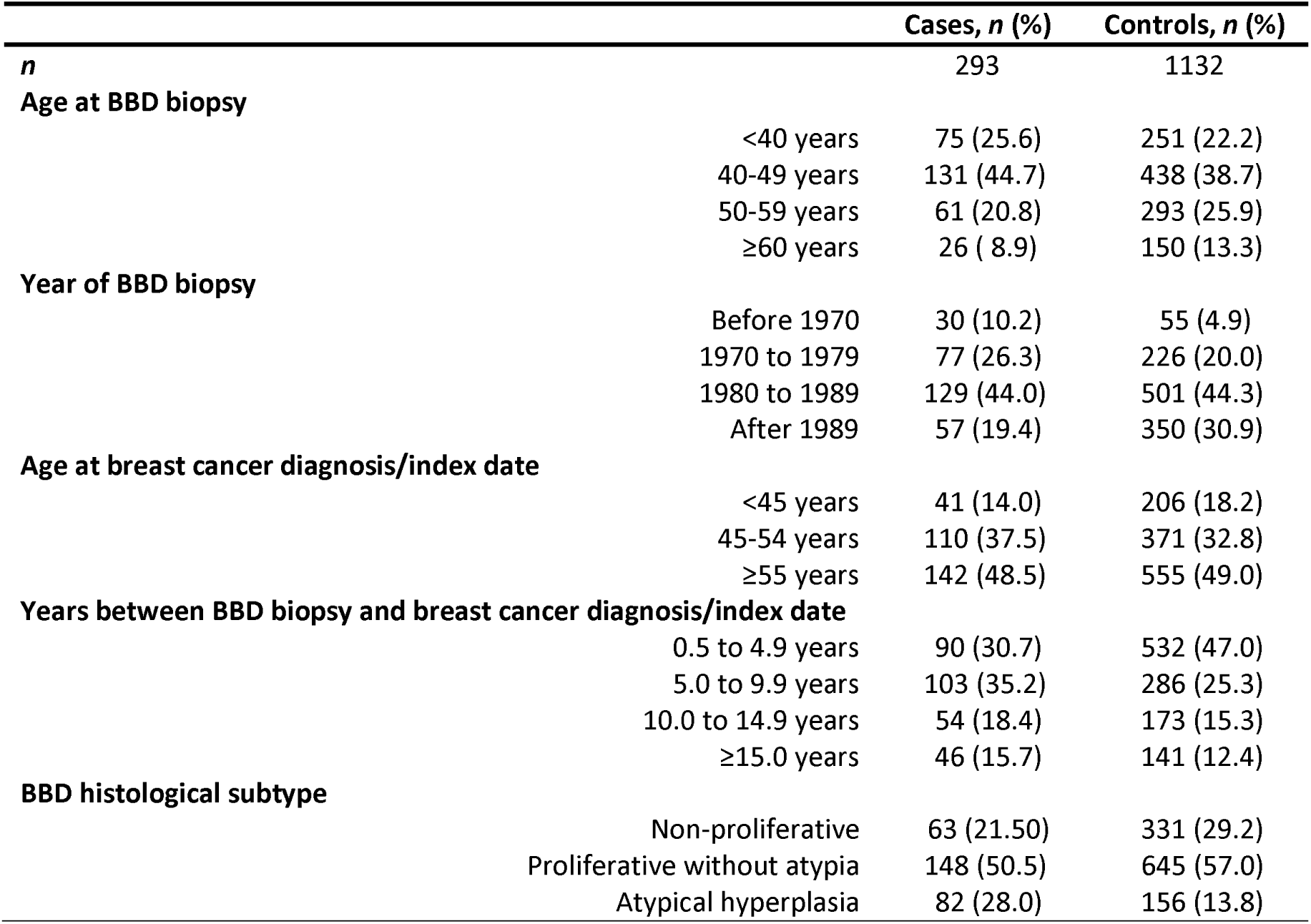
Benign breast disease (BBD) nested case-control study participants.

|  |  | Cases, *n* (%) | Controls, *n* (%) |
| --- | --- | --- | --- |
| *n* |  | 293 | 1132 |
| **Age at BBD biopsy** |  |  |  |
|  | <40 years | 75 (25.6) | 251 (22.2) |
|  | 40-49 years | 131 (44.7) | 438 (38.7) |
|  | 50-59 years | 61 (20.8) | 293 (25.9) |
|  | ≥60 years | 26 ( 8.9) | 150 (13.3) |
| **Year of BBD biopsy** |  |  |  |
|  | Before 1970 | 30 (10.2) | 55 (4.9) |
|  | 1970 to 1979 | 77 (26.3) | 226 (20.0) |
|  | 1980 to 1989 | 129 (44.0) | 501 (44.3) |
|  | After 1989 | 57 (19.4) | 350 (30.9) |
| **Age at breast cancer diagnosis/index date** |  |  |  |
|  | <45 years | 41 (14.0) | 206 (18.2) |
|  | 45-54 years | 110 (37.5) | 371 (32.8) |
|  | ≥55 years | 142 (48.5) | 555 (49.0) |
| **Years between BBD biopsy and breast cancer diagnosis/index date** |  |  |  |
|  | 0.5 to 4.9 years | 90 (30.7) | 532 (47.0) |
|  | 5.0 to 9.9 years | 103 (35.2) | 286 (25.3) |
|  | 10.0 to 14.9 years | 54 (18.4) | 173 (15.3) |
|  | ≥15.0 years | 46 (15.7) | 141 (12.4) |
| **BBD histological subtype** |  |  |  |
|  | Non-proliferative | 63 (21.50) | 331 (29.2) |
|  | Proliferative without atypia | 148 (50.5) | 645 (57.0) |
|  | Atypical hyperplasia | 82 (28.0) | 156 (13.8) |

### Preliminary assessment of breast tissue composition

Crude Wilcoxon tests compared cases and controls within each tissue region (Figure 3A). Cases have significantly more epithelium (*p*<0.001) and suggestively more fibrous stroma (*p* = 0.07) than controls. Controls have significantly more fat (*p*<0.001) than cases. When stratified by BBD histological subtypes, there were significant differences among cases or controls, or between cases and controls for each tissue region (epithelium *p*<0.001, fibrous stroma *p* = 0.02, fat *p*<0.001; Kruskal-Wallis tests). Dunn’s post-hoc analyses with Benjamini-Hochberg multiple testing adjustment revealed that among cases, those with proliferative without atypia subtype had significantly more epithelium compared to those with nonproliferative (adjusted p = 0.03) or proliferative with atypia subtype (adjusted p = 0.004; Figure 3B). Among controls, those with non-proliferative subtype had significantly less epithelium than those with proliferative without atypia (adjusted *p*< 0.001) or proliferative with atypia subtypes (adjusted p = 0.009; Figure 3B). When comparing between cases and controls, cases with nonproliferative or proliferative without atypia subtype have significantly more epithelium compared to controls (adjusted p = 0.02 and 0.002, respectively; Figure 3B).

**Figure 3.**
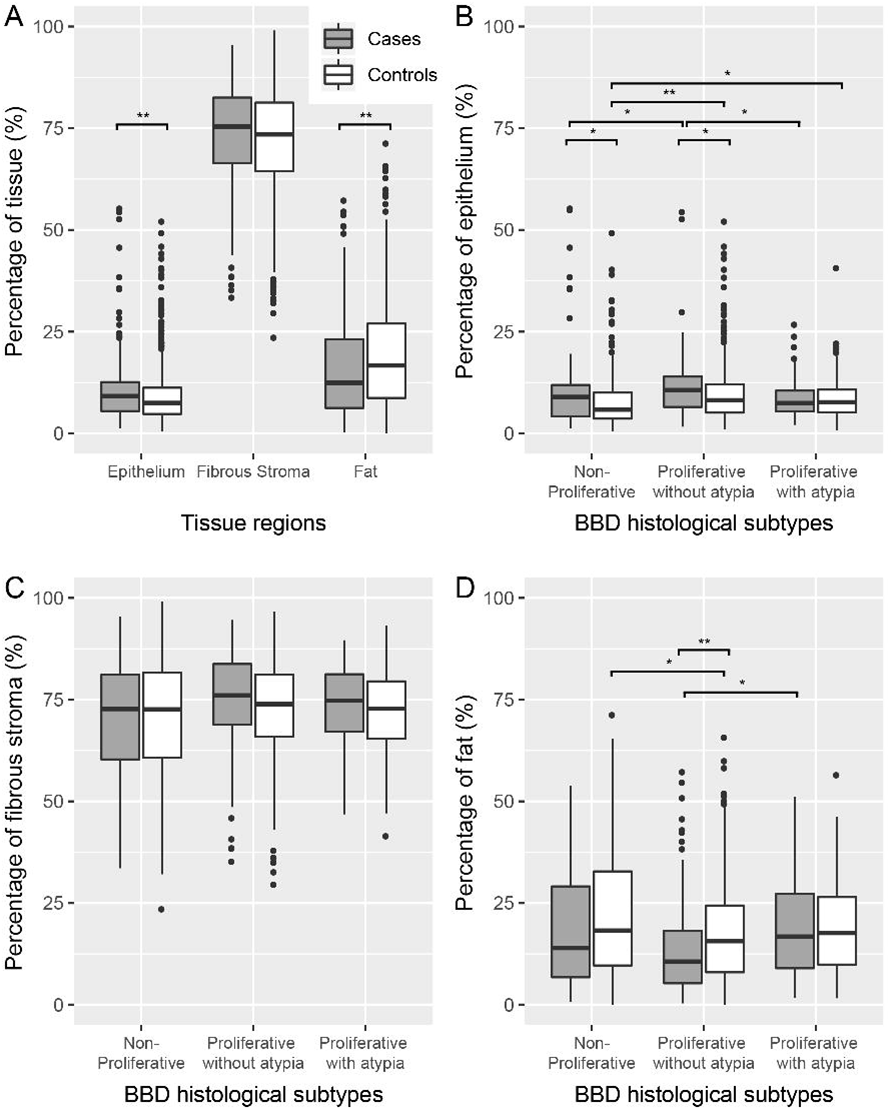
Boxplots display the amount of each tissue region (%) among cases and controls (A), and when stratified by benign breast disease (BBD) histological subtypes (B, C, D). * denotes *p*< 0.05 and ** denotes *p*< 0.001.

There was no pairwise difference in the amount of fibrous stroma among cases or controls, or between cases and controls within each BBD histological subtype in crude analyses (adjusted p>0.05 for all comparisons; Figure 3C). Cases with proliferative without atypia subtype have less fat than cases with non-proliferative (adjusted p = 0.05) or proliferative with atypia subtypes (adjusted p = 0.002; Figure 3D). Likewise, controls with proliferative without atypia subtype have less fat than controls with non-proliferative (adjusted p = 0.001) or proliferative with atypia subtypes (adjusted p = 0.09; Figure 3D). Within the proliferative without atypia subtype, cases have significantly less fat compared to controls (adjusted *p*< 0.001; Figure 3D).

### Age-adjusted tissue composition and breast cancer risk factors among controls

Table 2 displays the age-adjusted means (95% CI) and the ANCOVA p-values of the associations between BBD histological subtypes, body size, and reproductive breast cancer risk factors and the tissue composition among the controls. The relationship between BBD histological subtypes and percentages of epithelium and fat among controls remained significant in age-adjusted analyses (both *p*< 0.001). In contrast to crude analyses, controls with proliferative subtypes have significantly more fibrous stroma compared to controls with nonproliferative subtype after adjusting for age (*p*< 0.001).

**Table 2.**
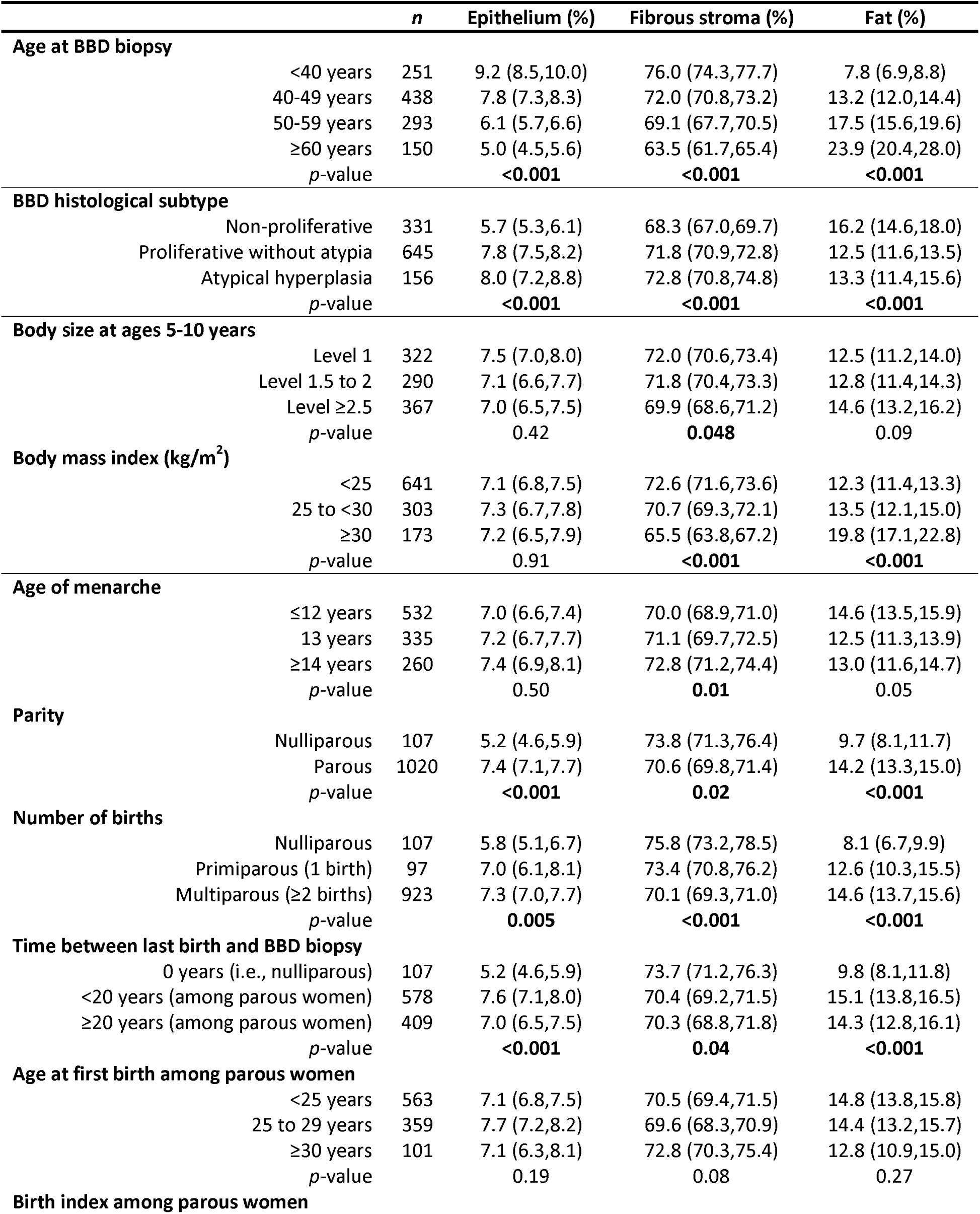
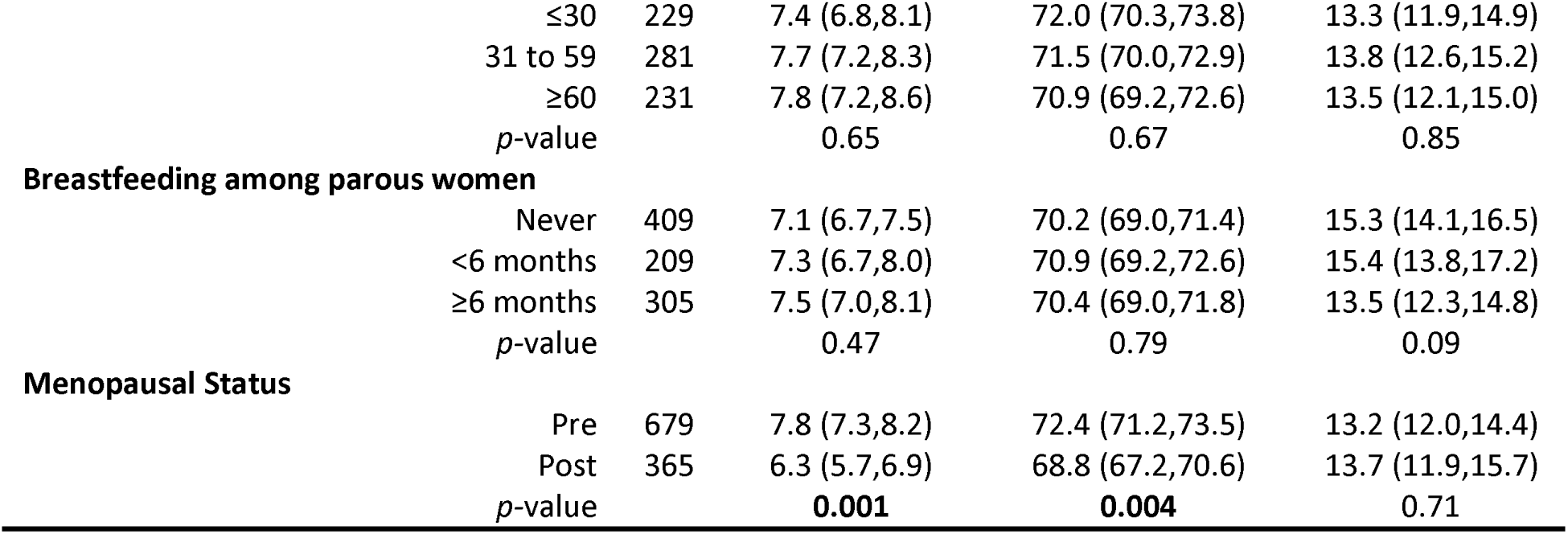
Tissue composition (%) and breast cancer risk factors among 1132 controls. Data presented for age are means (95% confidence interval). Data for other variables are presented as age-adjusted means (95% confidence interval); age was adjusted as a continuous variable.

|  | *n* | Epithelium (%) | Fibrous stroma (%) | Fat (%) |
| --- | --- | --- | --- | --- |
| **Age at BBD biopsy** |  |  |  |  |
| <40 years | 251 | 9.2 (8.5,10.0) | 76.0 (74.3,77.7) | 7.8 (6.9,8.8) |
| 40-49 years | 438 | 7.8 (7.3,8.3) | 72.0 (70.8,73.2) | 13.2 (12.0,14.4) |
| 50-59 years | 293 | 6.1 (5.7,6.6) | 69.1 (67.7,70.5) | 17.5 (15.6,19.6) |
| ≥60 years | 150 | 5.0 (4.5,5.6) | 63.5 (61.7,65.4) | 23.9 (20.4,28.0) |
| *p*-value |  | <0.001 | <0.001 | <0.001 |
| **BBD histological subtype** |  |  |  |  |
| Non-proliferative | 331 | 5.7 (5.3,6.1) | 68.3 (67.0,69.7) | 16.2 (14.6,18.0) |
| Proliferative without atypia | 645 | 7.8 (7.5,8.2) | 71.8 (70.9,72.8) | 12.5 (11.6,13.5) |
| Atypical hyperplasia | 156 | 8.0 (7.2,8.8) | 72.8 (70.8,74.8) | 13.3 (11.4,15.6) |
| *p*-value |  | <0.001 | <0.001 | <0.001 |
| **Body size at ages 5-10 years** |  |  |  |  |
| Level 1 | 322 | 7.5 (7.0,8.0) | 72.0 (70.6,73.4) | 12.5 (11.2,14.0) |
| Level 1.5 to 2 | 290 | 7.1 (6.6,7.7) | 71.8 (70.4,73.3) | 12.8 (11.4,14.3) |
| Level ≥2.5 | 367 | 7.0 (6.5,7.5) | 69.9 (68.6,71.2) | 14.6 (13.2,16.2) |
| *p*-value |  | 0.42 | 0.048 | 0.09 |
| **Body mass index (kg/m<sup>2</sup>)** |  |  |  |  |
| <25 | 641 | 7.1 (6.8,7.5) | 72.6 (71.6,73.6) | 12.3 (11.4,13.3) |
| 25 to <30 | 303 | 7.3 (6.7,7.8) | 70.7 (69.3,72.1) | 13.5 (12.1,15.0) |
| ≥30 | 173 | 7.2 (6.5,7.9) | 65.5 (63.8,67.2) | 19.8 (17.1,22.8) |
| *p*-value |  | 0.91 | <0.001 | <0.001 |
| **Age of menarche** |  |  |  |  |
| ≤12 years | 532 | 7.0 (6.6,7.4) | 70.0 (68.9,71.0) | 14.6 (13.5,15.9) |
| 13 years | 335 | 7.2 (6.7,7.7) | 71.1 (69.7,72.5) | 12.5 (11.3,13.9) |
| ≥14 years | 260 | 7.4 (6.9,8.1) | 72.8 (71.2,74.4) | 13.0 (11.6,14.7) |
| *p*-value |  | 0.50 | 0.01 | 0.05 |
| **Parity** |  |  |  |  |
| Nulliparous | 107 | 5.2 (4.6,5.9) | 73.8 (71.3,76.4) | 9.7 (8.1,11.7) |
| Parous | 1020 | 7.4 (7.1,7.7) | 70.6 (69.8,71.4) | 14.2 (13.3,15.0) |
| *p*-value |  | <0.001 | 0.02 | <0.001 |
| **Number of births** |  |  |  |  |
| Nulliparous | 107 | 5.8 (5.1,6.7) | 75.8 (73.2,78.5) | 8.1 (6.7,9.9) |
| Primiparous (1 birth) | 97 | 7.0 (6.1,8.1) | 73.4 (70.8,76.2) | 12.6 (10.3,15.5) |
| Multiparous (≥2 births) | 923 | 7.3 (7.0,7.7) | 70.1 (69.3,71.0) | 14.6 (13.7,15.6) |
| *p*-value |  | 0.005 | <0.001 | <0.001 |
| **Time between last birth and BBD biopsy** |  |  |  |  |
| 0 years (i.e., nulliparous) | 107 | 5.2 (4.6,5.9) | 73.7 (71.2,76.3) | 9.8 (8.1,11.8) |
| <20 years (among parous women) | 578 | 7.6 (7.1,8.0) | 70.4 (69.2,71.5) | 15.1 (13.8,16.5) |
| ≥20 years (among parous women) | 409 | 7.0 (6.5,7.5) | 70.3 (68.8,71.8) | 14.3 (12.8,16.1) |
| *p*-value |  | <0.001 | 0.04 | <0.001 |
| **Age at first birth among parous women** |  |  |  |  |
| <25 years | 563 | 7.1 (6.8,7.5) | 70.5 (69.4,71.5) | 14.8 (13.8,15.8) |
| 25 to 29 years | 359 | 7.7 (7.2,8.2) | 69.6 (68.3,70.9) | 14.4 (13.2,15.7) |
| ≥30 years | 101 | 7.1 (6.3,8.1) | 72.8 (70.3,75.4) | 12.8 (10.9,15.0) |
| *p*-value |  | 0.19 | 0.08 | 0.27 |
| **Birth index among parous women** |  |  |  |  |

|  |  |  |  |  |  |
| --- | --- | --- | --- | --- | --- |
|  | ≤30 | 229 | 7.4 (6.8,8.1) | 72.0 (70.3,73.8) | 13.3 (11.9,14.9) |
|  | 31 to 59 | 281 | 7.7 (7.2,8.3) | 71.5 (70.0,72.9) | 13.8 (12.6,15.2) |
|  | ≥60 | 231 | 7.8 (7.2,8.6) | 70.9 (69.2,72.6) | 13.5 (12.1,15.0) |
|  | *p*-value |  | 0.65 | 0.67 | 0.85 |
| **Breastfeeding among parous women** |  |  |  |  |  |
|  | Never | 409 | 7.1 (6.7,7.5) | 70.2 (69.0,71.4) | 15.3 (14.1,16.5) |
|  | <6 months | 209 | 7.3 (6.7,8.0) | 70.9 (69.2,72.6) | 15.4 (13.8,17.2) |
|  | ≥6 months | 305 | 7.5 (7.0,8.1) | 70.4 (69.0,71.8) | 13.5 (12.3,14.8) |
|  | *p*-value |  | 0.47 | 0.79 | 0.09 |
| **Menopausal Status** |  |  |  |  |  |
|  | Pre | 679 | 7.8 (7.3,8.2) | 72.4 (71.2,73.5) | 13.2 (12.0,14.4) |
|  | Post | 365 | 6.3 (5.7,6.9) | 68.8 (67.2,70.6) | 13.7 (11.9,15.7) |
|  | *p*-value |  | **0.001** | **0.004** | 0.71 |

Women with a larger childhood body size (Levels 1.5–2 and ≥2.5) had less fibrous stroma (*p* = 0.048) but suggestively more fat (*p* = 0.09) compared to women with body sizes of 1 or 1.5 to 2 at ages 5–10 years. Breast tissues of women with BMI ≥30 at the time of BBD biopsy had lower amount of fibrous stroma (*p*< 0.001) but higher amount of fat (*p*< 0.001) compared to women with lower BMI.

Parous women had more epithelium and fat, and less fibrous stroma compared to nulliparous women (all *p*< 0.05; Table 2). When parous women were further subdivided, women who had ≥2 births (multiparous) had more epithelium and fat, and less fibrous stroma than women who had one birth (primiparous) or nulliparous women (*p*< 0.05). When parous women were subdivided into women whose last birth was < 20 years or ≥20 years prior to BBD diagnosis, women who had their last birth <20 years had more epithelium and fat compared to nulliparous women and women who had their last birth ≥ 20 years prior to BBD diagnosis (*p*< 0.05). Post-menopausal women had less epithelium (*p* = 0.001) and fibrous stroma (*p* = 0.004) compared to pre-menopausal women (Table 2). The age of menarche positively correlated with the amount of fibrous stroma (*p* = 0.01). Age at first birth, birth index, and breastfeeding were not associated with breast tissue composition.

### Tissue composition and breast cancer risk

A higher proportion of epithelium was significantly associated with subsequent breast cancer risk when accounting for matching factors (OR = 1.53, 95% CI 1.04–2.27 comparing highest and lowest quartiles; *p-trend =* 0.02). Upon additional adjustment for BBD subtype, parity, menopausal status, and BMI, the association modestly attenuated but remained significantly elevated (OR = 1.39, 95% CI 0.91–2.14 comparing highest and lowest quartiles; *p-trend* = 0.047; Table 3). Neither the amount of fibrous stroma nor fat was associated with breast cancer risk in all three models (all *p-trend*>0.05; Table 3).

**Table 3.** The association between tissue composition and breast cancer risk was evaluated using unconditional logistic regression models to estimate odd ratios (ORs) and 95% confidence intervals (CI).

|  | Quartile 1 | Quartile 2 | Quartile 3 | Quartile 4 | *p-trend* |
| --- | --- | --- | --- | --- | --- |
| **Epithelium** |  |  |  |  |  |
| Cases/Controls, *n* | 56/283 | 65/283 | 68/283 | 104/283 |  |
| Quartile cutoff, % | <4.8 | ≥4.8 to <7.5 | ≥7.5 to <11.2 | ≥11.2 |  |
| Model 1 | Ref | 1.12 (0.76,1.67) | 1.12 (0.75,1.67) | 1.53 (1.04,2.27) | **0.02** |
| Model 2 | Ref | 0.95 (0.63,1.43) | 0.92 (0.61,1.39) | 1.36 (0.91,2.03) | 0.06 |
| Model 3 | Ref | 0.95 (0.61,1.49) | 0.95 (0.61,1.49) | 1.39 (0.91,2.14) | **0.047** |
| **Fibrous stroma** |  |  |  |  |  |
| Cases/Controls, *n* | 62/283 | 67/283 | 78/283 | 86/283 |  |
| Quartile cutoff, % | <64.5 | ≥64.5 to <73.5 | ≥73.5 to <81.3 | ≥81.3 |  |
| Model 1 | Ref | 0.98 (0.66,1.45) | 1.07 (0.73,1.57) | 1.20 (0.81,1.76) | 0.33 |
| Model 2 | Ref | 0.87 (0.58,1.30) | 0.96 (0.65,1.42) | 1.07 (0.72,1.59) | 0.65 |
| Model 3 | Ref | 0.78 (0.51,1.20) | 0.86 (0.56,1.31) | 0.93 (0.61,1.41) | 0.85 |
| **Fat** |  |  |  |  |  |
| Cases/Controls, *n* | 102/283 | 80/283 | 49/283 | 62/283 |  |
| Quartile cutoff, % | <8.7 | ≥8.7 to <16.7 | ≥16.7 to <27.0 | ≥27.0 |  |
| Model 1 | Ref | 0.81 (0.57,1.15) | 0.55 (0.36,0.81) | 0.75 (0.50,1.12) | 0.11 |
| Model 2 | Ref | 0.81 (0.56,1.15) | 0.55 (0.36,0.82) | 0.83 (0.55,1.25) | 0.27 |
| Model 3 | Ref | 0.83 (0.58,1.21) | 0.56 (0.36,0.85) | 0.93 (0.59,1.45) | 0.52 |

Each tissue region was categorized into quartiles as defined by the distribution among the controls. Model 1 adjusted for matching factors. Model 2 adjusted for matching factors and BBD histological subtypes. Model 3 adjusted for matching factors, BBD histological subtypes, parity, menopausal status, and BMI. The median value for each quartile was included as a continuous variable in the unconditional logistic regression for Model 1, 2, and 3, to obtain the *p-trend* value (Wald test).

Additional analyses were conducted, stratifying by BBD, parity, menopausal status, or BMI. Within the BBD proliferative without atypia subtype, women with epithelium in the fourth quartile had higher breast cancer risk compared to women in the first quartile (adjusted OR = 1.92, 95% CI 1.11–3.40; *p-trend* = 0.01; Supplementary Table 2). Polytomous logistic regression models assessed the association between tissue regions and breast cancer risk defined by tumor ER expression, and in general demonstrated no heterogeneity. Fat was associated with lower breast cancer risk among ER-positive women in the crude model 1 (second versus first tertile: OR = 0.62, 95% CI 0.42–0.92; third versus first tertile: OR = 0.62, 95% CI 0.41–0.95; *p-trend* = 0.04; Supplementary Table 3).

To understand the substitution effects, further analyses were conducted using each tissue region as a continuous variable per 10% change and with two of the three tissue regions in the model (Supplementary Table 4). Similar to Table 3 where tissue regions were analyzed using quartiles, the association between epithelium as a continuous variable and breast cancer risk remained the strongest. Every 10% increase in the amount of epithelium was significantly associated with increased breast cancer risk in fully adjusted models, irrespective of whether it is substituted for fibrous stroma (adjusted OR = 1.30, 95% CI 1.05–1.61) or fat tissue (adjusted OR = 1.26, 95% CI 1.03–1.54; Supplementary Table 4).

### Morphometric signature associated with subsequent breast cancer risk

The morphometric signature developed using elastic net regularized regression consisted of four features in the epithelium: the kurtosis of delaunay edge length (fourth moment of a graph-based feature which measures the spatial density (interconnectedness) of nuclei found in each epithelial region), the kurtosis of euler number (fourth moment of a topological feature which measures the number of holes in an epithelial region), mean perimeter of epithelial regions, and the minimum blue channel pixel value of nuclei found in epithelial regions. The area under the receiver operator curve (AUC ROC) of the model was 0.61 (optimal *λ* = 0.08). When evaluated on the test set of 570 women, the AUC ROC was 0.51. Due to the poor AUC ROC of the test set, the association of the signature score with breast cancer was not further evaluated.

## Discussion

The identification of new biomarkers may improve breast cancer risk prediction. We developed a deep-learning based computational pathology method to segment BBD histopathological images into epithelial, fibrous stroma, and fat regions; calculate tissue composition; and extract morphometric features. We applied our method to the BBD breast cancer NCC study within the NHS/NHSII to determine the association of the tissue composition with breast cancer risk factors and breast cancer risk. Among controls, BBD subtypes, parity, and number of births were significantly associated with all three tissue regions; select tissue regions were associated with body size at ages 5–10 years, BMI, age of menarche, and menopausal status. Women whose breast tissues had higher percentages of epithelium had significantly increased risk of breast cancer compared to women with lower percentages, especially among women with proliferative without atypia subtype of BBD. We also extracted computer-derived morphometric features from each tissue region but were unable to construct a robust breast cancer morphometric signature. Our study showed that the percentage of epithelium may be utilized as a potential biomarker of breast cancer risk.

BBD and breast cancer originate from TDLUs. The epithelium captured by our computational method was all-encompassing—normal TDLUs, TDLUs with proliferative or metaplastic changes, and various types of BBD lesions. Hence, our study was the first to demonstrate a direct quantitative relationship between the percentage of epithelium and subsequent breast cancer risk in women diagnosed with BBD. Our findings also supported the long-held hypothesis that elevated cellular mass increases risk of cancer^53^. Some lesion types within the proliferative without atypia subtype are highly cellular. For example, adenosis is characterized by increased number of enlarged lobules while radial scar is characterized by a central area of fibroelastotic change from which epithelial structures with or without associated hyperplasia or metaplasia radiate. This explained why when stratified by BBD subtype, the association of the percentage of epithelium and breast cancer risk remained significant among women within the proliferative without atypia of BBD.

The associations of age-adjusted breast tissue composition and breast cancer risk factors among controls provided histopathological evidence to support epidemiological studies, mainly by demonstrating the link between breast tissue cellularity and cancer risk^53^. Our work suggests that risk factors have different influences on the percentages of epithelium and fibrous stroma. Gertig *et al*. evaluated the proportion of epithelium and fibrous stroma in 300 BBD women who did not develop breast cancer^54^. We and Gertig *et al*. showed that breast tissues associated with the non-proliferative subtype of BBD were less cellular (i.e., lower epithelium and fibrous stroma but higher fat percentages) than proliferative with or without atypia subtypes, thus partly explaining why women with the non-proliferative subtype have lower breast cancer risk compared to proliferative subtypes^33,55–58^.

Adiposity during childhood or in young adults is inversely associated with breast cancer risk^59–61^. Body adiposity is correlated with the amount of fat tissue in the breast when evaluated using percentage mammographic density (i.e., proportion of dense (epithelium and fibrous stroma) to non-dense tissues (fat))^62,63^. As such, percentage mammographic density is also inversely associated with breast cancer risk. In 153 normal breast tissue samples obtained via core-biopsy, Gabrielson *et al*. observed significant inverse associations of BMI with percentages of epithelium and stroma^64^. However, our study and the study by Gertig *et al*^54^ were conducted using more participants and only observed a significant inverse association between BMI and proportion of stroma. Nevertheless, these three studies provided histologic evidence to partially explain the differential breast cancer risk by adiposity—breast tissues of women with a larger childhood body size or younger women with BMI ≥30 have lower overall cellularity (i.e., epithelium and fibrous stroma) and thus are less dense compared to women with a leaner childhood body size or women with lower BMI, respectively.

Parity had the strongest influence on breast tissue composition among the reproductive risk factors investigated in our study. Gertig *et al*. and Gabrielson *et al*. observed more epithelium and less fibrous stroma in parous women compared to nulliparous women^54,64^. Our observation of higher percentages of epithelium and fat, and lower fibrous stroma percentages in multiparous women who had a live birth within the last 20 years was similar to other studies that observed less TDLU involution in parous versus nulliparous women^24,65^; supported epidemiological reports of increased breast cancer risk in parous women who had a live birth within the last 5 to 24 years compared to nulliparous women^66^; and highlighted the extensive stroma remodeling in mammary glands during pregnancy to accommodate expanding epithelium^67^. The correlation between age of menarche and proportion of stroma reported by us and others^54,64^ is in line with higher percent breast density in young women who had later ages of menarche^68^. The null associations between age of first birth and length of breastfeeding with breast tissue composition agreed with Gertig *et al*.^54^ while Gabrielson *et al*.^64^ found an association between percentage of epithelium and length of breastfeeding (β = 0.03±0.01, p = 0.02), but not percentage of stroma (β = 0.43±0.26, p = 0.10). Using a different computational pathology method that specifically measures normal TDLUs, we also did not find an association between length of breastfeeding and TDLU involution^24^. Older women have less dense breasts than younger women, with the greatest change in density occurring during the menopause years^69^. Indeed, we and Gertig *et al*.^54^ reported that post-menopausal women had less epithelium and fibrous stroma compared to pre-menopausal women. However, this was not observed by Gabrielson *et al*.^64^, possibly due to low power.

Computer-derived morphometric signatures have shown potential as prognostic or diagnostic biomarkers^15,16,70^. We did not identify a breast cancer morphometric signature from BBD WSIs. Morphometric feature data are typically noisy. In an effort to reduce signal noise, we attempted unsuccessfully to create a breast cancer signature within each BBD histopathological subtype due to low power. There were more features than the number of samples. Extracting and combining morphometric features from different types of epithelium may have excessively diluted meaningful signals. Using the median metric^15^, a common method of aggregating morphometric features across all WSIs, may not optimal for this dataset. There is no gold standard method for feature aggregation and this remains an active area of research by medical image analysis groups. Future work can include improving methods for morphometric feature aggregation or create specific breast cancer morphometric signatures for each type of BBD lesion.

The strengths of our study include the application of a computer pathology method to assess breast tissue composition in a large nested case-control study with rich data on breast cancer risk factors^3,4,26,27,32,34^. BBD samples underwent centralized pathology review and breast cancer cases were confirmed through review of medical records. Some limitations of our study include being underpowered to evaluate the association of breast composition and ER negative breast cancer, breast cancer molecular subtypes^71,72^, or mammographic density^73,74^ as mammogram data were only available for 105 women (7.8%) in this study. Our findings were also limited to White women, the predominant race of the NHS/NHSII participants. Dysfunctional epithelial-stroma interactions in the breast have been implicated in breast carcinogenesis^75^, however, our current study was not designed to investigate epithelium-stroma interactions.

In conclusion, we found that BBD histopathological subtypes, anthropometric, and selective reproductive risk factors were associated with breast tissue composition. Higher percentages of epithelium were associated with increased risk of breast cancer, specifically among women with the proliferative without atypia subtype of BBD. No morphometric signature was associated with subsequent breast cancer. Future work can include incorporation of the percentage of epithelium into risk assessment models as well as explore end-to-end deep learning breast cancer prediction models.

## Data Availability

The data that support the findings of this study are available from the Nurses' Health Studies, however they are not publicly available. Investigators interested in using the data can request access, and feasibility will be discussed at an investigators meeting. Limits are not placed on scientific questions or methods, and there is no requirement for co-authorship. Additional data sharing information and policy details can be accessed at http://www.nurseshealthstudy.org/researchers.
The source code is available on GitHub

https://github.com/avellal14/BBD_Pipeline

## Acknowledgements

We thank the participants and staff of the Nurses’ Health Study and Nurses’ Health Study II for their valuable contributions as well as the following state cancer registries for their help: AL, AZ, AR, CA, CO, CT, DE, FL, GA, ID, IL, IN, IA, KY, LA, ME, MD, MA, MI, NE, NH, NJ, NY, NC, ND, OH, OK, OR, PA, RI, SC, TN, TX, VA, WA, WY. The authors assume full responsibility for analyses and interpretation of these data.

## Conflict of interest statement

The authors declare no conflicts of interest.

## Funding

This work was supported by the National Institute of Health/National Cancer Institute R21CA187642 (RMT), R01CA175080 (RMT), R01CA240341 (RMT, YJH), UM1CA186107 (AHE), and U01 CA176726 (AHE), Susan G. Komen for the Cure IIR13264020 (RMT), the Klarman Family Foundation (YJH), BIDMC High School Summer Research Program (ADV), and the Deep Learning for Medical Image Analysis research program by Netherlands Organization for Scientific Research and Philips Research P15–26 (MV).

## Authors’ contributions

Conceived and designed the study: RMT YJH KS. Data analysis: YJH ADV KS MV KHK RMT. Epidemiological data collection: RMT KHK. Breast pathology expertise: GMB LCC SJS JLC. Computational method and data acquisition: ADV KS ALS MEP YJH. All authors contributed to the writing and reviewing of the manuscript.

---

**Supplementary Figure 1.**
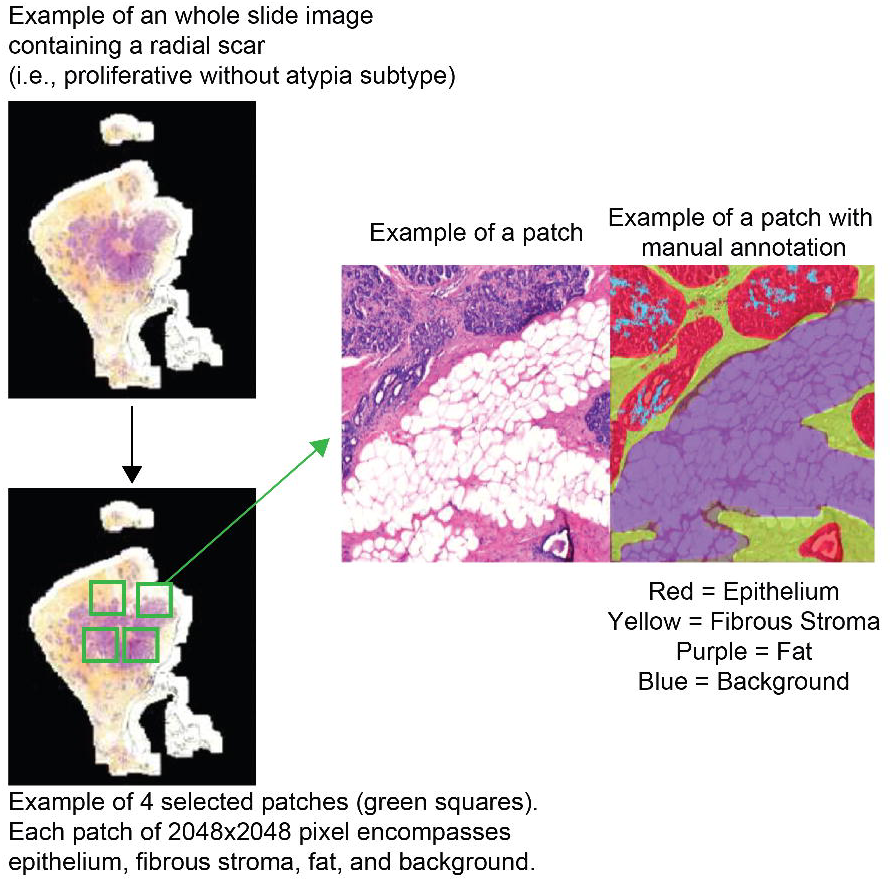
A schematic diagram showing how four patches were selected on a whole slide image for manual annotation. The dark areas of the whole slide image correspond to areas of the slide that do not contain tissue and were thus not scanned by the scanner.

**Supplementary Figure 2.**
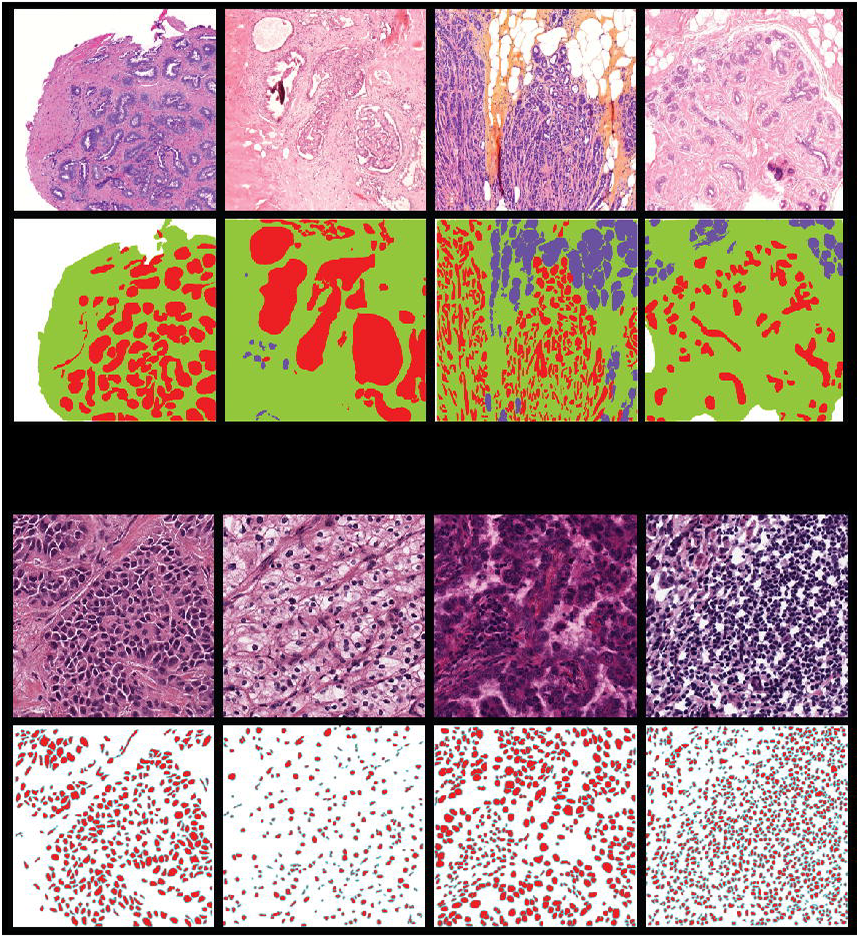
Example patches used to train the tissue segmentation and nuclei detection networks. The top row in each section display the original patches, the bottom row contains the corresponding manually annotated ground truths. Images display a broad spectrum of staining colors and intensities. **(A)** Examples of tissue segmentation training patches from the Nurses’ Health Studies benign breast disease incidence study. In our ground truth annotations, white represents background, green represents fibrous stroma, red is epithelium, and purple is fat. **(B)** Examples of nuclei detection training patches from invasive breast cancer cases from The Cancer Genome Atlas. In the ground truth annotations, white represents background, red is nucleus, and cyan is nuclei membrane border.

## Notes

### Competing Interest Statement

The authors have declared no competing interest.

### Author Declarations

The study protocol was approved by the institutional review boards of the Brigham and Women's Hospital and Harvard T.H. Chan School of Public Health, and those of participating registries as required.

